# SARS-CoV-2 infection and Pregnancy outcome: A cross-sectional study from Eastern U.P. Population, India

**DOI:** 10.1101/2024.02.15.24302882

**Authors:** Sangeeta Rai, Ashish Ashish, Shivani Mishra, Bhupendra Kumar, Kusum Kusum, Royana Singh

## Abstract

Pregnant women with coronavirus infection are at a higher risk for severe diseases. In the present study, we evaluated and compared clinical characteristics and outcomes in pregnancy of normal females and females with SARS-CoV-2 infection. Our study was a cross-sectional study. The pregnant females were examined, their blood samples were taken for Covid Panel (D-Dimer, Ferritin, IL-6, CRP, PCT (Procalcitonin)); and oral-nasal swabs were taken for SARS-CoV-2 infection. Both SARS-CoV-2 positive and negative (control) females were followed up every trimester for any complication related to pregnancy. We found that females suffering from SARS-CoV-2 infection had reduced gestation periods, and had higher percentage of caesarean and pre-term delivery than SARS-CoV-2 negative females. Based on our findings, it appears that there exist close associations between SARS-CoV-2 infection in pregnant females and increased risk of reduced gestation periods, and spontaneous caesarean and pre-term delivery. However, more studies are still needed to validate present findings.

## 1. Introduction

Soon after the worldwide outbreak of COVID-19 infection in year 2020, WHO verified the epidemic as a global public health emergency of International Concern (Sohrabi et al., 2020). Currently, the clinical researches on novel Coronavirus (SARS-CoV-2) are limited. The multiple variants of SARS-CoV-2 are also circulating globally (Harvey et al., 2021). It is still unknown whether these variants have clinical impacts on the pregnant women. Although previous studies including pregnant women and the coronavirus family diseases, like severe acute respiratory syndrome coronavirus (SARS-Co-V) and the Middle East Respiratory Syndrome coronavirus (MERS-CoV) have reported close associations of these diseases with severe complications during pregnancy, such as miscarriage, preeclampsia, fetal growth restriction, preterm birth, prelabor rupture of membranes, and maternal deaths (M et al., 2020; Schwartz & Graham, 2020).

Consequently, the pregnant women are at a higher risk of developing SARS-Co-V-2 infection and other associated diseases, if neglected (Knight et al., 2020). Studies have shown that pregnant women are predominantly susceptible to respiratory pathogens and severe pneumonia (Ellington, 2021; Kayem et al., 2020). The immediate physiological changes in the immune and cardiopulmonary systems of pregnant women after coronavirus infection, i.e. elevated diaphragm, increased rate of oxygen consumption, and edema of respiratory tract mucosa, make the females at the higher risk of developing hypoxia (C et al., 2020). Thus, there is an urgent need to generate the population-specific data of pregnant women, so as to explore the social-epidemiological and clinical determinants of the outcomes of COVID-19 in them (Ellington, 2021).

Currently, in India, the cases of COVID-19 are on a constant rise. The Ministry of Health and Family Welfare (MoHFW), Government of India, has reported 3,30,58,843 total confirmed cases with 4,41,042 deaths, as on7^th^September 2021. Out of these, the state of Uttar Pradesh has alone contributed about 17,09,457 cases with about 22,861 deaths due to COVID-19 (*MoHFW* | *Home*, n.d.). Hence there is an urgent need to pay special attention for this state in planning the strategies for combating the SARS-CoV-2 infection, especially in the vulnerable population group, which include the pregnant women. In the present study, we have evaluated and compared the clinical characteristics and the outcomes in pregnancy of normal females and the females with SARS-CoV-2 infection. We anticipate that the findings will help us in understanding the close association between SARS-CoV-2 infection and pregnancy, and to develop future strategies to control SARS-CoV-2 infection in pregnant females.

## 2. Materials and Methods

Our study is a cross-sectional study done in pregnant females (n=36) during the period of February 2021 to June 2021 All the pregnant females coming to Sir Sunderlal Hospital, Banaras Hindu University (BHU), Varanasi (India) were triaged. Based on their health history the females were examined following the standard precautions for SARS-CoV-2 infection after taking their proper consent. The study was approved by ethical committee of the Institution, Banaras Hindu University (No.Dean/2021/EC/2762).Their blood samples were sent to Institute of Medical Sciences (IMS), BHU, Varanasi for Covid Panel (D-Dimer, Ferritin, IL-6, CRP, PCT (Procalcitonin); and their oral and nasal swabs were taken for examining SARS-CoV-2 infection and sent to MRU -lab, IMS, BHU, Varanasi. The pregnant females tested positive were sent to COVID-19 ward for further management. The females who were found negative were considered as control. Both the positive (n=19) and control (n=17) females were followed up every trimester for any complication related to pregnancy and their data were properly recorded for further statistical analysis.

## 3. Statistical analyses

The distributions of data sets obtained in this study were checked for normality using Kolmogorov-Smirnoff test. Means were separated using Tukey’s test when data were normally distributed and variances were homogeneous (Bartlett’s test for equal variances). The obtained data on maternal age, maternal systolic and diastolic blood pressures, maternal body temperature, maternal haemoglobin content, gestation period, foetal heart rate and neonatal weight of both the SARS-CoV-2 positive and SARS-CoV-2 negative females were subjected to unpaired t-test, followed by Tukey’s *post-hoc* comparison of means.

However, the percent data comparing:(i) normal versus caesarean delivery, (ii) full-term versus pre-term delivery, (iii) immediate and delayed cord clamping, and (iv) oxytocin administered versus no oxytocin administered delivery, in both SARS-CoV-2 positive and SARS-CoV-2 negative females were subjected to two variable χ^2^-test. All statistical analyses were performed using MINITAB 16 (Minitab Inc., State College, Pennsylvania, United States of America) on PC.

## 4. Results

In the present study, all the subjects were young pregnant females of age group 26-27 years. They were either normal females without SARS-CoV-2 infection (n=17) or the females having SARS-CoV-2 infection (n=19). The physiological parameters revealed that irrespective of SARS-CoV-2 infection, the systolic and diastolic blood pressures of the females were 118-134 mmHg and 74-78 mmHg, respectively. The haemoglobin content, foetal heart rate and neonatal weights were 10-11 gm/dL, 136-145 per minute and 2-2.5kg, respectively. Unpaired t-test did not detect significant difference in the maternal systolic and diastolic blood pressures, maternal haemoglobin content, foetal heart rate and neonatal weight of SARS-CoV-2 positive and SARS-CoV-2 negative females. However, the values of gestation period differed significantly between SARS-CoV-2 positive and SARS-CoV-2 negative females. Tukey’s *post-hoc* comparison of means revealed that females suffering from SARS-CoV-2 infection had reduced gestation periods than those without SARS-CoV-2 infection.

The χ^2^-values further exposed significant differences in normal versus caesarean delivery, full-term versus pre-term delivery, and oxytocin administered versus no oxytocin administered delivery of SARS-CoV-2 positive and SARS-CoV-2 negative females. Comparison of means revealed that SARS-CoV-2 negative females had higher percentage of oxytocin administered (92.86%) normal (23.53%) and full term (61.11%) delivery than SARS-CoV-2 positive females (84.21%, 5.26% and 61.11%, respectively). In contrast, SARS-CoV-2 positive females had higher percentage of caesarean (94.74%) and pre-term (38.89%) delivery than SARS-CoV-2 negative females (76.47% and 11.76%, respectively).

## Discussion

In the present study, females suffering from SARS-CoV-2 infection had reduced gestation periods, and had higher percentage of caesarean and pre-term delivery than SARS-CoV-2 negative females. The reduced gestation periods and pre-term delivery may solely be due to maternal respiratory compromise, as reported earlier in pregnant women with confirmed SARS-CoV-2 infection in the United Kingdom (Knight et al., 2020). In their study, Knight et al. (2020) also reported that 12% of the women admitted with SARS-CoV-2 infection were delivered preterm, and almost 60% of women gave birth by caesarean section. Moreover, one in twenty of the babies had a positive test for SARS-CoV-2, and half had infection diagnosed on samples taken at less than 12 hours after birth.

The higher rates of caesarean births by SARS-CoV-2 infected females in our study also indicate towards the maternal compromise due to SARS-CoV-2 infection. In a meta-analysis study involving 1316 pregnant women, the investigators found that the cesarean deliveries occurred among three-fourths of the pregnant women infected with corona viruses (Diriba et al., 2020). Regarding the perinatal outcome, the fetal distress and neonatal asphyxia were the most commonly reported abnormalities in infants.

Many previous studies have also reported that the SARS-CoV-2 infection during pregnancy may cause complications to both the mother and the fetus (Lei et al., 2020; Villar et al., 2021). Such complications include elevated blood pressure, preeclampsia, preterm and premature rupture of placental membranes, restricted fetal growth, and/or miscarriage. A large number of CoV-infected pregnant women with severe cases were admitted to the intensive care unit which even resulted in the death of some of the mothers. Contrary to our findings, many researchers did not report any association between SARS-CoV-2 infection during pregnancy and the increased risk of frequent abortions and spontaneous preterm births (Wan et al., 2020; Yan et al., 2020).

Based on our findings, it appears that there exist close associations between SARS-CoV-2 infection in pregnant females and the increased risk of reduced gestation periods, and spontaneous caesarean and pre-term delivery. However, more studies are still needed considering the population groups of other regions of India to further validate the present findings.

## Author Contribution

RS, AA and SR contributed to the data collection and interpretation and the initial draft of the manuscript. BK carried out the statistical analysis and contributed to the draft and revision of the manuscript. SM AND KK contributed to the data collection and analysis. BK contributed to the data collection and analysis. YQ design of the study and the patient recruitment and critically revised the manuscript. All authors provided and approved for the final manuscript

## Data Availability

All data produced in the present work are contained in the manuscript

## Acknowledgement

This research was sponsored by Multi-Disciplinary Research Units (MRUs) and DST, a grant by ICMR-Department of Health Research [Grant No: 6004].

---

**Table 1.**
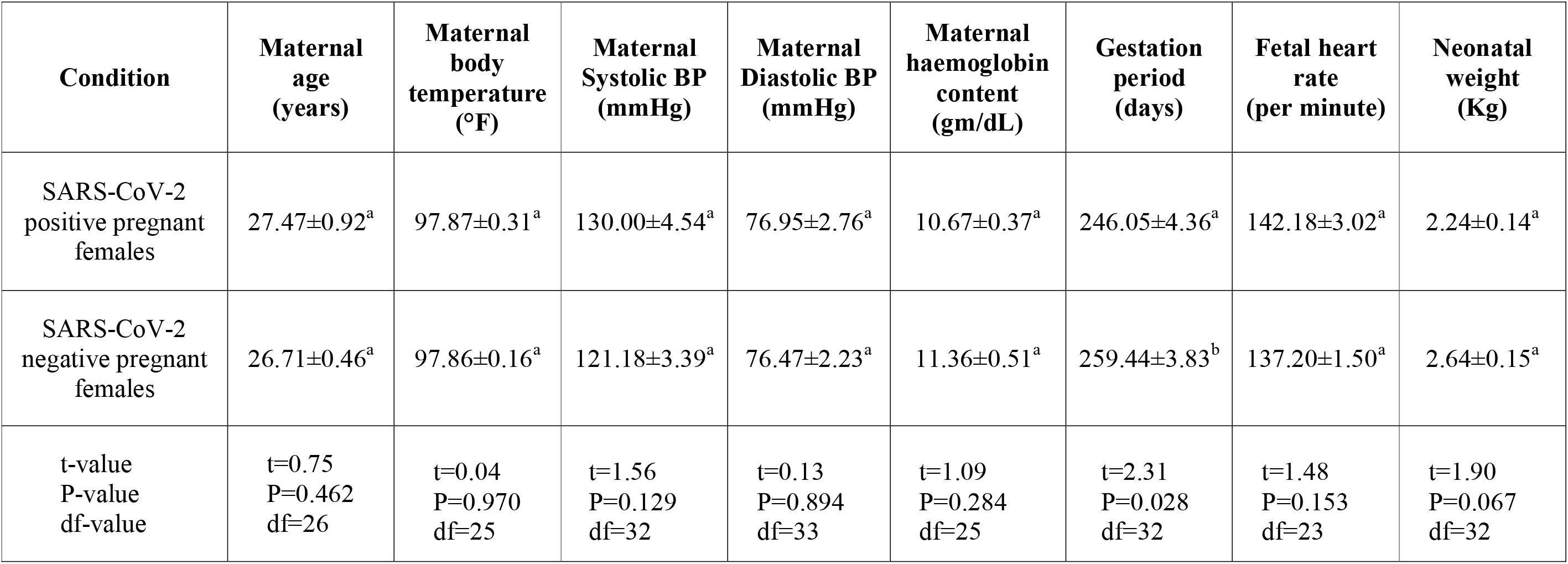
Effect of various physiological parameters of pregnant females with and without SARS-CoV-2 infection (Values are Mean±SE; tvalues significant at P<0.05; small letters represent comparison of means between normal and SARS-CoV-2 infection pregnant females).

| Condition | Maternal age (years) | Maternal body temperature (°F) | Maternal Systolic BP (mmHg) | Maternal Diastolic BP (mmHg) | Maternal haemoglobin content (gm/dL) | Gestation period (days) | Fetal heart rate (per minute) | Neonatal weight (Kg) |
| --- | --- | --- | --- | --- | --- | --- | --- | --- |
| SARS-CoV-2 positive pregnant females | 27.47±0.92<sup>a</sup> | 97.87±0.31<sup>a</sup> | 130.00±4.54<sup>a</sup> | 76.95±2.76<sup>a</sup> | 10.67±0.37<sup>a</sup> | 246.05±4.36<sup>a</sup> | 142.18±3.02<sup>a</sup> | 2.24±0.14<sup>a</sup> |
| SARS-CoV-2 negative pregnant females | 26.71±0.46<sup>a</sup> | 97.86±0.16<sup>a</sup> | 121.18±3.39<sup>a</sup> | 76.47±2.23<sup>a</sup> | 11.36±0.51<sup>a</sup> | 259.44±3.83<sup>b</sup> | 137.20±1.50<sup>a</sup> | 2.64±0.15<sup>a</sup> |
| t-value<br>P-value<br>df-value | t=0.75<br>P=0.462<br>df=26 | t=0.04<br>P=0.970<br>df=25 | t=1.56<br>P=0.129<br>df=32 | t=0.13<br>P=0.894<br>df=33 | t=1.09<br>P=0.284<br>df=25 | t=2.31<br>P=0.028<br>df=32 | t=1.48<br>P=0.153<br>df=23 | t=1.90<br>P=0.067<br>df=32 |

**Table 2.** Chi-square table comparing the parameters related to child birth in females with and without SARS-CoV-2 infection (Values are percent values; χ^2^-value significant at P<0.05; small letters represent comparison of means between normal and SARS-CoV-2 infection females).

| Parameters | SARS-CoV-2 positive pregnant females | SARS-CoV-2 negative pregnant females | $\chi^2$-value; P-value; df |
| --- | --- | --- | --- |
| Normal delivery (%) | 5.26<sup>a</sup> | 23.53<sup>b</sup> | $\chi^2=14.559$; $P < 0.0001$; df=1 |
| Caesarean delivery (%) | 94.74<sup>a</sup> | 76.47<sup>a</sup> |  |
| Full-term delivery (%) | 61.11<sup>a</sup> | 88.24<sup>b</sup> | $\chi^2=19.187$; $P < 0.0001$; df=1 |
| Pre-term delivery (%) | 38.89<sup>b</sup> | 11.76<sup>a</sup> |  |
| Immediate cord clamping (%) | 13.33<sup>a</sup> | 7.14<sup>a</sup> | $\chi^2=2.000$; $P=0.157$; df=1 |
| Delayed cord clamping (%) | 86.67<sup>a</sup> | 92.86<sup>a</sup> |  |
| Oxytocin administered (%) | 84.21<sup>a</sup> | 92.86<sup>b</sup> | $\chi^2=3.979$; $P=0.046$; df=1 |
| No oxytocin administered (%) | 15.79<sup>b</sup> | 7.14<sup>a</sup> |  |

